# Enisamium reduces influenza virus shedding and improves patient recovery by inhibiting viral RNA polymerase activity

**DOI:** 10.1101/2020.12.23.20247569

**Authors:** Tatiana G. Zubkova, Aartjan J.W. te Velthuis, Megan Shaw, Andrew Mehle, David Boltz, Norbert Gmeinwieser, Holger Stammer, Jens Milde, Lutz Müller, Victor Margitich

**Author notes:** Corresponding authors: Dr. Regenold GmbH, Zoellinplatz 4, 79410 Badenweiler, Germany, Division of Virology, Department of Pathology, Addenbrooke’s Hospital, University of Cambridge, Hills Road, CB2 2QQ, United Kingdom; Fax: +44 1223, 333346. Department of Medical Biosciences, University of the Western Cape, Robert Sobukwe Road, Bellville 7535, South Africa (Megan Shaw).

## Abstract

Infections with respiratory viruses constitute a huge burden on our health and economy. Antivirals against some respiratory viruses are available, but further options are urgently needed. Enisamium iodide (laboratory code FAV00A, trade name Amizon®) is an antiviral marketed in countries of the Commonwealth of Independent States for the treatment of viral respiratory infections, but its clinical efficacy and mode of action are not well understood. Here, we investigated the efficacy of FAV00A in patients aged between 18-60 years with confirmed influenza and other viral respiratory infections. FAV00A treatment resulted in reduced influenza virus shedding (at day 3, 71.2% in FAV00A group tested negative versus 25.0% in placebo group, p < 0.0001), faster patient recovery (at day 14, 93.9% in FAV00A group had recovered versus 32.5 % in placebo group, p < 0.0001), and reduced disease symptoms compared to placebo (from 9.6 ± 0.7 to 4.6 ± 0.9 score points in FAV00A group versus 9.7 ± 1.1 to 5.6 ± 1.1 score points in placebo group, p < 0.0001). Using mass-spectrometry, and cell-based and cell-free viral RNA synthesis assays, we identified a hydroxylated metabolite of FAV00A, VR17-04. VR17-04 is capable of inhibiting influenza virus RNA synthesis and present in plasma of patients treated with FAV00A. VR-17-04 inhibits the activity of the influenza virus RNA polymerase more potently than its parent compound. Overall, these results suggest that FAV00A is metabolized in humans to an inhibitor of the influenza virus RNA polymerase that reduces viral shedding and improves patient recovery in influenza patients.

Clinical data are available on ClincalTrials.gov under NCT04682444.

## Introduction

Infections with respiratory viruses, such as influenza A virus (IAV), are the cause of morbidity and mortality in humans, and a source of serious medical and socio-economic problems for human health worldwide. Of additional concern is the simultaneous circulation of several respiratory viruses in the human population, including influenza B virus (IBV), adenoviruses (ADV), parainfluenza viruses (PIV), respiratory syncytial virus (RSV), coronaviruses (CoV), and various IAV subtypes (e.g., H3N2 and H1N1). All these viruses have the potential to spread rapidly among the human population, become resistant to antivirals, cause severe disease, and be associated with secondary complications (1). Moreover, novel respiratory viruses may emerge from animal reservoirs, such as wild birds or bats, and cause a pandemic when no or little pre-existing immunity exists in the human population. Examples of recent outbreaks are the SARS-CoV-2 pandemic virus in 2019-2020 and the H1N1 pandemic IAV strain in 2009-2010. Together, these characteristics make respiratory viruses a major threat to human health and extraordinarily difficult target for the development of preventive and therapeutic mitigation measures.

Seasonal IAV and IBV infections can be prevented by inactivated or live attenuated vaccines. Vaccines have also been approved for emerging highly pathogenic IAV H5N1 or H7N9 strains, and pandemic SARS-CoV-2. However, the efficacy of these vaccines is dependent upon antigenic similarity between the vaccine strain and the circulating viruses, and potentially diminished in high-risk groups (2, 3). Furthermore, the lead-time to produce new vaccines is long. Currently, no vaccines exist against human seasonal CoV or RSV infections, although vaccines are in development (4-6).

Drug-based antiviral therapies are an important mitigation strategy against respiratory virus infections. In the case of IAV infections, drugs are available that target the viral neuraminidase protein (e.g., oseltamivir, zanamivir, peramivir and lainamivir), the IAV RNA-dependent RNA polymerase (e.g., favipiravir) or the endonuclease domain of PA protein (baloxavir marboxil) (7, 8). Emergence of resistant viruses has been reported for most of these antivirals (9, 10) and research into alternative antiviral therapies with a different mode of action is needed.

Enisamium is an isonicotinic acid derivative. The iodide salt of enisamium (FAV00A), formerly named carbabenzpyride, is marketed as Amizon® in 11 countries, including in former Soviet Union countries and Mongolia, for the prophylaxis and treatment of influenza and other viral diseases in adults and children (11). Previous studies indicate that FAV00A inhibits IAV and IBV replication in cell culture (12).

IAVs and IBV are negative-sense RNA viruses whose 14 kb genome consists of eight segments of single-stranded viral RNA (vRNA). The viral RNA-dependent RNA polymerase copies the vRNA into a replicative intermediate called the complementary RNA (cRNA) during viral replication, or into capped and polyadenylated viral messenger RNA (mRNA) during viral transcription (13). The cRNA serves as a template for the production of new vRNA molecules. vRNA and cRNA molecules are both replicated in the context of ribonucleoproteins (RNPs), which consist of a copy of the RNA polymerase complex bound to the 5’ and 3’ ends of a genome segment and a helical coil of nucleoproteins (NPs) that are bound to the rest of the vRNA or cRNA (13). The IAV and IBV RNA-dependent RNA polymerases are composed of three subunits: polymerase basic 1 (PB1) protein, PB2 protein, and polymerase acidic (PA) protein. All three protein subunits are essential for replication and transcription of the viral RNA genome segments (13).

Here we present a comprehensive analysis of the efficacy of FAV00A against IAV *in vitro*, and IAV and IBV infections in patients. We find that FAV00A is effective in reducing viral shedding and improving patient recovery in influenza infected patients. Subsequent mass-spectroscopy analysis of sera from FAV00A-treated patients revealed the presence of a hydroxylated metabolite of FAV00A, VR17-04. Using cell-free RNA polymerase activity experiments, we show that the metabolite VR17-04 is a more potent inhibitor of influenza A viral RNA synthesis than FAV00A. Overall, these results suggest that FAV00A is effective in reducing virus shedding and improving recovery of influenza patients, and its mode of action is inhibition of the influenza virus RNA polymerase most likely via the active metabolite VR17-04.

## Materials and methods

### Ethics of clinical study

To investigate efficacy and tolerability of FAV00A in patients infected with respiratory viruses, a prospective, single-blinded, placebo-controlled, single-center clinical trial was performed. The study was conducted in accordance with the Declaration of Helsinki, ICH-GCP and the national laws and regulations in the Research Institute of Influenza of the North-West Branch of the Russian Academy of Medical Sciences (NWB RAMS), St. Petersburg, Russian Federation, between 2009 and 2010. The study was approved by the Ethics Commission of the Research Institute of Influenza of the NWB RAMS (Extract of report No 28 of 09 April 2009) after review of the clinical trial protocol, Investigator’s Brochure, patient information with informed consent, case report forms, insurance policy, and the qualifications of the investigator. Patients provided a written informed consent prior to study participation.

### Sample size of clinical study

A sample size calculation was performed to design the clinical trial. The maximal difference between pre- and post-treatment parameters that was considered not significant was set to 0.25. Thus, a sample size of 30 patients in both the FAV00A- and placebo-treated groups would be needed to demonstrate an efficacy of FAV00A with a statistical power of 90% (bilateral) and a confidence level of 5%. We further estimated that about 15% of the patients would withdraw or be withdrawn (see exclusion criteria below) from the study prior to its completion. Using Fisher’s exact test for subsequent analyses, we estimated that a starting cohort of 50 patients per group would be sufficient to achieve the statistical power stated above.

### Patient cohort and treatment during the clinical study

Male and female patients aged between 18 and 60 years with viral respiratory infections were eligible for participation in the clinical study. The diagnosis of viral respiratory infection was based on axillary body temperature ≥ 37.2°C, the presence of at least one symptom of respiratory tract affection (rhinitis, pharyngitis, laryngitis, tracheitis, bronchitis, cough), and one general infection symptom (weakness, malaise, myalgia, headache, fever, decreased appetite). In addition, diagnosis was confirmed by viral antigen immunofluorescence testing of nasal swabs. Patients with any acute organ dysfunction, serious chronic disease, pregnancy, or breastfeeding obligations were excluded from clinical trial participation. Patients were recruited by advertisements, randomized into two groups, and treated with either FAV00A (500 mg three times daily) or matching placebo for 7 days. Daily dosage and duration of treatment were in accordance with the summary of product characteristics of FAV00A. The use of immunomodulatory drugs, systemic sympathomimetics, antipyretics, analgesics, and antibiotics was prohibited during the clinical trial, unless it was required for intervention. Endpoints of the study were the dynamics of clinical signs by investigating the disappearance of signs of rhinitis and the decrease of body temperature, interferon status and reduction of the time to recovery.

### Data collection in clinical study

During the course of the clinical trial, 4 visits were scheduled for each patient: a baseline visit (day 0) and three follow-up visits 3, 7 and 14 days thereafter. Objective disease symptoms were assessed by the investigator and subjective symptoms by the patients during each visit. The objective symptoms comprised fever (i.e., body temperature ≥37.2°C), pharyngeal hyperemia, conjunctival infection, enlarged lymph nodes, abnormal arterial blood pressure, and auscultation findings (lung and heart). The subjective symptoms included weakness, myalgia, headache, increased body temperature (i.e., subjective severity rating), chills, sore throat and cough. All patients were monitored for adverse events throughout the course of the clinical trial.

A score system was used to assess patient health. Objective symptoms were scores as follows: normal or abnormal blood pressure was counted 0 or 4 score points; lung auscultation was counted 0 for vesicular breath sound and wheezing or crepitation were scored 2 or 4 points, respectively; clear and rhythmic heart sounds were each scored 0 points, whereas noisy and arrhythmic heart sounds were scored 2 points each. The subjective symptoms were assessed using a 4-point Likert scale, ranging from 1 (absent) to 4 (severe). After patient assessment, a sum score was calculated. These scores ranged from 4 to 28 (minimum/maximum) for objective symptoms and from 7 to 28 score point for subjective symptoms.

In addition to scoring patient’s health, information about the number of days since day 0 without routine activities due to respiratory infection and overall treatment efficacy (complete, significant or moderate improve, no significant change, worsened) was recorded by the patient and investigator. Finally, nasal swabs were collected from the patients on days 0, 3 and 7 to determine viral antigen levels by viral antigen immunofluorescence testing.

### Statistical analysis

Descriptive summary statistics were used for analyses of the clinical results. Patients with any after baseline efficacy and/or safety data were included in the respective analysis set. Continuous variables were described by mean, median, standard deviation (SD), minimum, as well as maximum, and categorical variables by absolute numbers and percentages. Interferential analyses were performed as primary comparison for categorical endpoints evaluated in a by-visit-manner. For categorical non-binary endpoints, separate van Elteren test stratified by baseline status was applied per visit or overall, whatever was applicable. If approximation of the chi-square distribution was not given, Fisher’s exact test was used for testing without adjustment for baseline status. For continuous endpoints collected over time, a mixed model repeated measurement was used with considered endpoint as dependent variable, baseline as covariate and treatment, visit and time multiplied by visit as fixed effects. Subgroup analyses by antigen type (IAV or IBV, IAV alone, IBV alone, ADV alone, a combination influenza/adenovirus, and others) were performed for selected efficacy parameters as described in the results section.

### Metabolite screening during pharmacokinetic study

To investigate the metabolization of FAV00A in humans, plasma samples were obtained from participants in a study aimed to investigate the influence of food intake on the pharmacokinetics of a single dose of FAV00A and conducted in the Vienna General Hospital at the Medical University of Vienna, Vienna, in 2009/2010. The study was performed in accordance with the Declaration of Helsinki, ICH-GCP and Austrian clinical trial law (EudraCT No. 2009-015382-32), and approved by the Ethics Commission of the Medical University Vienna and the General Hospital Vienna (reference no. 868/2009, dated 27 November 2009) after review of the clinical trial protocol, Investigator’s Brochure, patient information with informed consent, insurance policy, and qualification of the investigator. Patients were recruited from the database of the study site. During the study, 24 healthy volunteers were randomized and assigned to two groups in a cross-over design: fasting/fed, fed/fasting. Plasma samples (3 ml, K_3_EDTA Greiner-one Biotubes) were taken pre-dose (0 h) and following dosing at 0.25, 0.5, 0.75, 1, 1.25, 1.5, 1.75, 2, 2.5, 3, 3.5, 4, 6, 8, 12, 16, 24 and 48 h and stored at -20° C. For the metabolite screening 12 plasma samples from the fasting group were used.

To perform the metabolite analysis, the fixed volume of 50 μl (100 μl for pre-dose) of all 12 subjects at different time-points were pooled [pre-dose, P1 (0.5 + 1 + 1.5), P2 (2 + 2.5 + 3), P3 (3.5 + 4 + 6), P4 (8 + 12 + 16), P5 (24 + 48)] samples. Next, 1 mL of each plasma pool was precipitated with 3 mL of ethanol/acetonitrile (50 : 50, v/v) at room temperature. Plasma samples were centrifuged for 20 min at 3360 *g* at 8° C and the supernatant removed. Each precipitate was washed twice with 3 mL ethanol/acetonitrile (50 : 50, v/v) and centrifuged again for 20 min at 3360 *g* at 8° C. The supernatants of each wash step were subsequently combined and centrifuged again to collect any remaining precipitate. The final precipitate was dried under a stream of nitrogen and reconstituted in 100 μl water/acetonitrile (90 : 10, v/v). A 10 μl aliquot was further diluted with 40 μl of water/acetonitrile (90 : 10, v/v) and 10 μl of this solution was injected onto the high-performance liquid chromatography (HPLC) column.

LC separation of metabolites was achieved on a Thermo Hypersil Keystone, HYPERCARB (2.1□by 150□mm, 5 μm) column utilizing a binary gradient of 0.1 % formic acid in water (solvent A), and 0.1 % formic acid in water/acetonitrile (10 : 90, v/v, solvent B). The gradient schedule was programmed at 95 : 5 (A : B) for 0 to 1.0□min; 0 : 100 for 30□min, and 95 : 5 for 40□min. The LC flow rate was 200□µl/min. Each sample extract was analyzed by reverse-phase liquid chromatography with full scan high-resolution mass spectrometry detection. Experiments were carried out in full scan (positive ion mode, m/z range 120 – 1000, Scan-Event 1).

For selected pools, experiments were also carried out in MS/MS mode (Scan-Event 2 – 4). MS^n^ data were recorded in the data dependent mode at single mass resolution. In the MS^n^ experiments, the instrument operated using an inclusion list of all candidates, detected in full scan. Whenever a compound with the same m/z values as the compounds of the inclusion list was detected, the instrument automatically switched to MS^n^ and collected the fragmentation pattern. For the linear Ion trap methods, the experiments were carried out in full scan (positive ion mode, m/z range 120 – 1000, Scan-Event 1) and in MS/MS mode (Scan-Event 2 – 4). For LTQ-Orbitrap-Method (accurate mass measurements) the experiments were carried out in full scan mode (positive ion mode, m/z range 120 – 1000) with a resolution of 60’000 at m/z 400.

### Cells, FAV00A, influenza virus infections

Human epithelial lung carcinoma (A549), human rhabdomyosarcoma (RD), human hepatocellular carcinoma (HepG2), human embryonic kidney (HEK 293T), and Madin-Darby canine kidney (MDCK) cells were purchased from the American Type Culture Collection (ATCC, Manassas, VA). Human epithelial adenocarcinoma (Caco-2) cells were kindly provided by Dr. Martin Walsh (Mount Sinai School of Medicine, New York, NY). All cells were cultured at 37 °C, 5% CO2 in DMEM supplemented with 10% fetal calf serum (FBS) and 1% pen/strep. The Caco-2 and HepG2 cells were grown on collagen-coated plates. FAV00A was synthesized by Farmak and dissolved in DMSO. Favipiravir triphosphate (T-705-TP) was purchased by Santa-Cruz Biotechnology and dissolved in water.

A549 cells were preincubated with varying concentrations of FAV00A in Minimal Essential Medium (MEM) containing 0.5% FCS at 37 °C for 1 h. Cells were next infected with influenza A/WSN/33 (H1N1) virus at a multiplicity of infection (MOI) of 0.01 in MEM containing 0.5% FCS for 1 h. Following infection, the inoculum was removed and replaced with varying concentrations of FAV00A in MEM containing 0.5% FCS and incubated for 48 h. Plaque assays were performed on MDCK cells in MEM containing 0.5% FCS with a 1% agarose overlay and grown for 48 h at 37 °C. Uninfected cells were treated with compounds in parallel for assessment of toxicity using a CellTiter-Blue kit (Promega).

Caco-2, RD and HepG2 cells were pretreated with FAV00A for 6 h prior to infection with influenza A/WSN/33 (H1N1) virus and reapplied after infection. Supernatants were collected at 48 h post-infection and titers determined by immunofluorescence staining in 96-well plates as follows. Serial dilutions of the supernatants were made and used to infect monolayers of MDCK cells in a 96-well plate. After 1 h incubation, the virus was removed and the cells were washed with PBS, then incubated in post-infection media (DMEM, 0.1% FBS, 0.3% BSA, 1% pen/strep, 1 mg/ml TPCK trypsin) for 6 h at 37 °C. Cells were fixed in 4% paraformaldehyde, washed 3 times in PBS, and permeabilized in 0.5% Triton X-100. Infected cells were stained with a mouse monoclonal antibody against influenza virus NP (HT103, generated at Mount Sinai School of Medicine, New York, NY), and detected with a goat anti-mouse secondary antibody labeled with AlexFluor 488 (Invitrogen). The number of infected cells was quantified on an imaging plate cytometer (Celigo, Nexcelom), and used to determine the titer (pfu/ml) based on a standard curve from a known titer virus stock. Uninfected cells were treated with compounds in parallel for assessment of toxicity.

### Measurement of viral RNA levels in mini-genome assay

HEK 293T cells were transfected with plasmids expressing the subunits of the influenza A/WSN/33 (H1N1) virus RNA polymerase (pcDNA3-PB1, pcDNA3-PB2, pcDNA3-PA), the viral nucleoprotein (pcDNA3-NP) and a viral RNA template based on segment 5 (NP-encoding genome segment) (14, 15). As negative control, PB1 active site mutant (PB1a) was used in which the active site SDD motif was mutated to SAA (14). FAV00A was added to the cell culture medium 15 min after transfection. Twenty-four h after transfection, cells were washed in PBS and the total RNA extracted using Trizol and isopropanol precipitation (16). 5S rRNA, and IAV vRNA and mRNA steady state levels were subsequently analyzed by radioactive primer extension and 6% denaturing PAGE as described previously (16). Phosphorimaging was performed on an FLA7000 Typhoon Scanner (GE Healthcare). GFP experiments were performed through transfection of pcDNA3-eGFP and measured using a SpectraMax plate reader.

### Measurement of RNA polymerase activity in a cell-free system

The subunits of the A/WSN/33 (H1N1) influenza virus RNA polymerase with a TAP-tag on PB2 (pcDNA3-PB1, pcDNA3-PB2-TAP, pcDNA3-PA) were expressed in HEK 293T cells and purified as heterotrimeric complex using IgG sepharose chromatography (16). Next, 0.5 mM CTP, 0.5 mM UTP, 0.05 mM ATP, 0.01 mM GTP, 0.5 mM ApG, 0.001 mM [α-^32^P]GTP, T-705 triphosphate, FAV00A and VR17-04 at the concentrations indicated, and 0.5 µM 5’ and 3’ vRNA promoter strands were added to the purified RNA polymerase. After incubation at 30 °C for 15 min, samples were analyzed by 20% denaturing PAGE and autoradiography (14, 16).

## Results

### Patient cohort was predominantly infected with influenza A and B viruses

A total of 137 patients were recruited and screened for respiratory virus infections, and 100 were randomized for treatment with FAV00A (n = 60) or the placebo control (n = 40) (Fig. 1A). Age and gender distributions were similar between the two groups (Fig. 1B). All patients completed the trial as per protocol and no dropouts occurred. At the start of the clinical trial, the patients reported 1.6 ± 0.5 (mean ± SD) days without routine activity due to respiratory infection. Antigen testing showed that the patients treated with FAV00A or placebo were positive for IAV (43.0%), ADV (16.0%), a combination of IAV and ADV (14.0%), or IBV (12.0%). An additional 8.0% of patients tested positive for PIV or RSV, whereas 7% of the patients (5% in the FAV00A group and 10% in the placebo group) were tested negative for IAV, IBV, ADV, PIV or RSV antigens. Overall, 69% of patients were infected with influenza A and/or B viruses. Both the FAV00A- and placebo-treated groups had a similar distribution of respiratory viruses (Fig. 1C). Throughout the study, there was no deviation from the scheduled medication intake, with a mean treatment duration of 6.8 ± 0.8 days. Topical decongestants were used for a shorter duration of treatment and by a lower proportion of patients treated with FAV00A, when compared to the placebo group (23.3%, n = 14 vs. 75%, n = 30). Expectorants were used by all patients in both treatment groups, but the majority of the FAV00A-treated patients used the drug for 7 days compared to 14 days for the placebo-treated patients.

**Figure 1.**
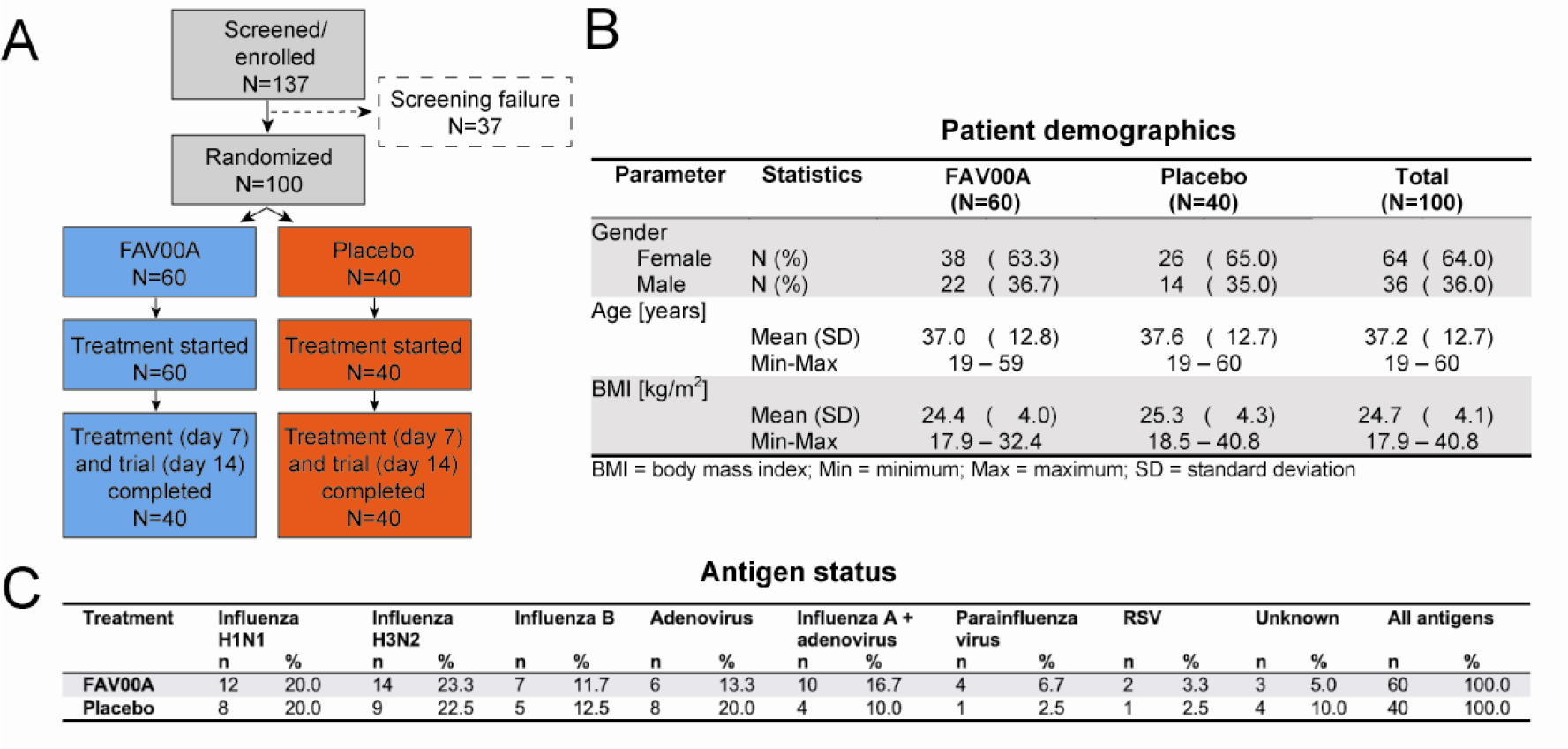
Patient enrollment, demographics and antigen status. A) Schematic of patient recruitment, randomization and treatment. B) Demographics of FAV00A- and placebo-treated patient groups. C) Frequency of virus antigen detection in nasal swabs of patients treated with FAV00A or placebo by immunofluorescence staining.

### FAV00A treatment reduces viral antigen levels and improves activity in patients with viral respiratory disease

Antigen testing on day 3, the first visit after initiation of treatment showed that 71.2% (n = 42) of the patients treated with FAV00A and 25.0% (n = 10) of the placebo group were negative for the above-mentioned respiratory virus antigens (p < 0.0001). On day 7, the second visit after initiation of treatment, all patients treated with FAV00A tested negative for respiratory virus antigens, while 82.5% (n = 33) of the placebo control patients tested negative (p = 0.0013) (Fig. 2A). The proportion of patients that tested antigen negative on days 3 and 7 after initiation of treatment was significantly higher (p < 0.0001) in the FAV00A-treated group (70.7%, n = 41) compared to placebo-treated group (25.0%, n = 10). A subgroup analysis indicated a similar positive effect of the FAV00A treatment when compared to the placebo for patients infected with IAV or IBV (75.0%, n = 24 and 22.7%, n = 5, respectively) or ADV (71.4%, n = 5 and 12.5%, n = 1, respectively). Analysis of patient behavior during the clinical study showed that the proportion of patients with no or just one day without routine activities after the start of the treatment was higher in the FAV00A-treated group compared to the placebo group. In the latter group the majority of patients was not able to perform routine activities for two or more days (Fig. 2B). These group differences were highly significant (p < 0.0001).

**Figure 2.**
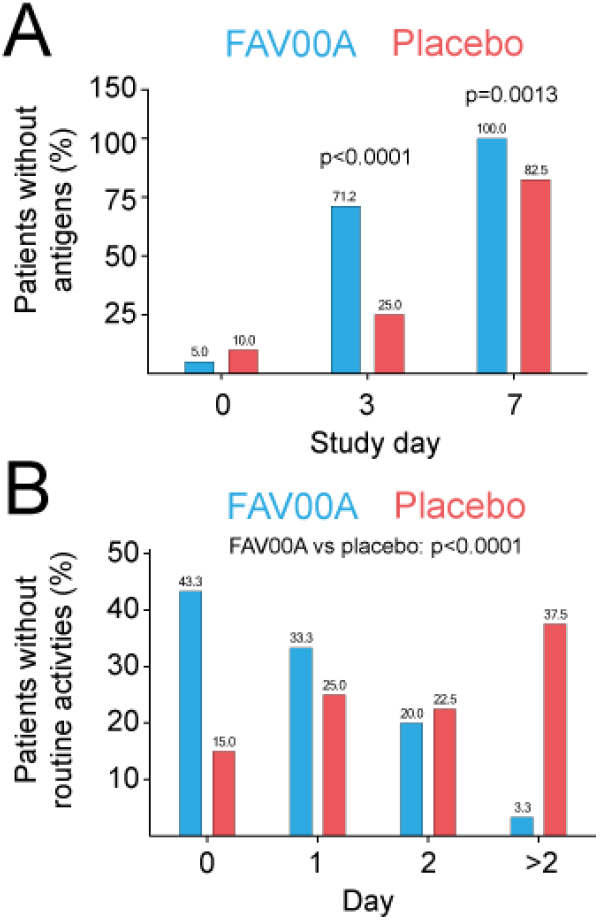
FAV00A treatment reduces viral antigen levels and improves patient activity. A) Patients in whom virus antigens were not detected by immunofluorescence staining of nasal swabs (%). B) Patients without routine activities (%). P values were determined by Fisher’s exact test.

### FAV00A treatment reduces objective symptoms in patients with viral respiratory disease

Analysis of the objective symptoms fever, pharyngeal hyperemia and abnormal lung auscultation contributed mainly to the objective symptom score at the start of the clinical trial on day 0. There was no difference between the objective symptom scores between the two groups on day 0 (9.6 ± 0.7 for FAV00A group and 9.7 ± 1.1 for placebo). A low proportion of trial participants presented conjunctival infection (22.0%) and enlarged lymph nodes (1.0%) and none had abnormal auscultation of the heart or arterial blood pressure at enrollment. The decrease in the objective symptom score from day 0 till day 14 was statistically significant (p < 0.0001) in the FAV00A-treated group (from 9.6 ± 0.7 to 4.6 ± 0.9 score points) when compared to the placebo group (from 9.7 ± 1.1 to 5.6 ± 1.1 score points). Absence of all objective symptoms was observed in both patient groups starting from day 7 (Fig. 3A) and a significant increase in the absence of all 3 symptoms was observed in the FAV00A-treated patients on day 14 (Fig. 3A; p < 0.0001). Fever, pharyngeal hyperemia as well as lung auscultation were similar in both treatment groups on day 0, and pharyngeal hyperemia as well as lung auscultation resolved significantly faster in the FAV00A group compared to the placebo group (Fig. 3B-C). Together, these observations strongly suggest that FAV00A treatment leads to a faster and highly significant reduction in objective clinical symptoms associated with respiratory virus infections.

**Figure 3.**
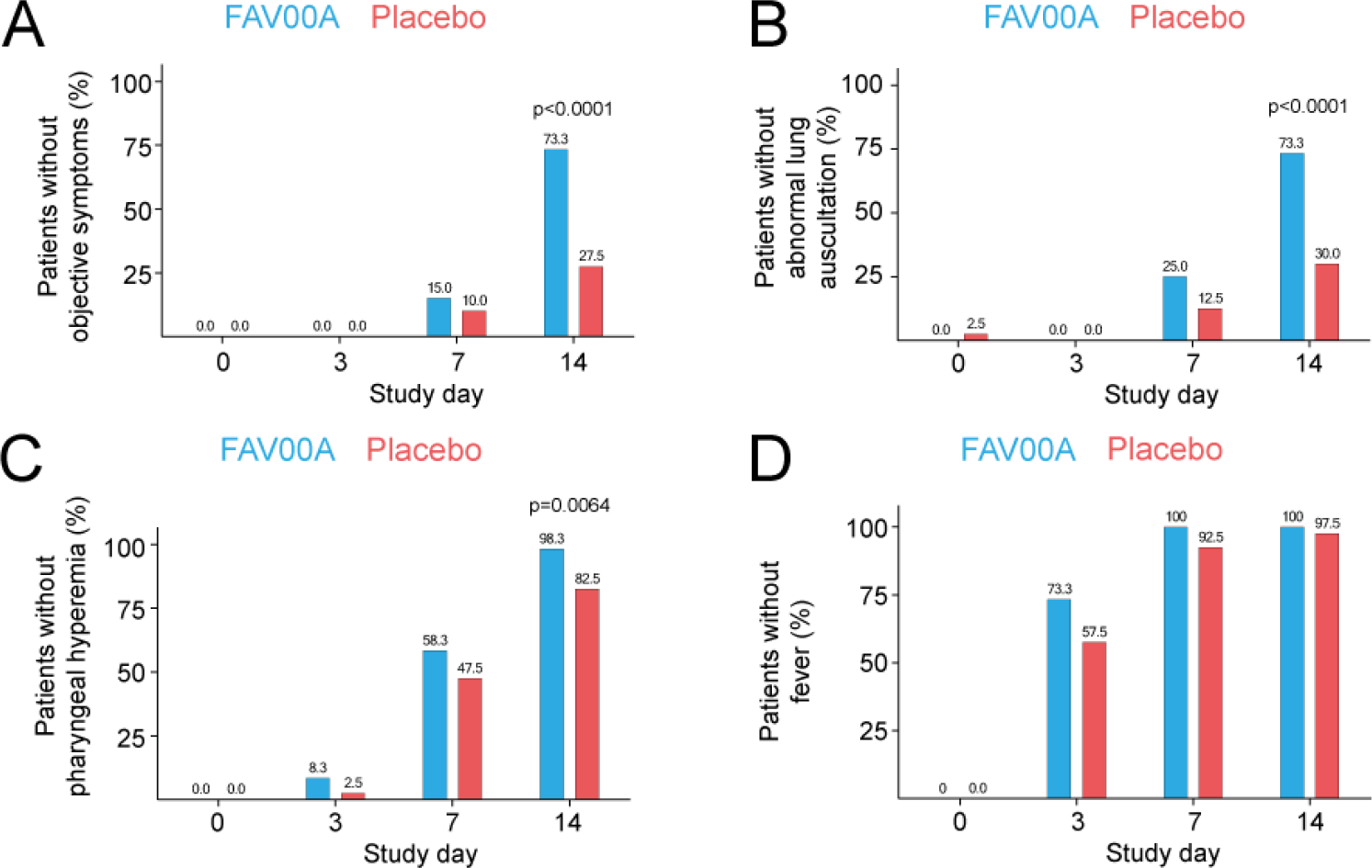
FAV00A treatment reduces objective symptoms in patients with viral respiratory disease. A) Patients without objective symptoms at different visit days (%). B) Patients without abnormal breath sounds at different visit days (%). C) Patients without pharyngeal hyperemia at different visit days (%). D) Patients without fever at different visit days (%). P values were determined by Fisher’s exact test.

### FAV00A treatment reduces subjective symptoms in patients with viral respiratory disease

The subjective symptoms weakness, headache, increased perceived body temperature, sore throat, and cough were present in all patients on day 0 and contributed most to the mean subjective symptom score. Less than half of the patients reported myalgia (46.0%, n = 46) and just a few participants had chills (7.0%, n = 7) at day 0. However, the subjective symptoms were predominantly of mild intensity and almost absent on day 3 in both groups, meaning that they had limited impact on the score.

Analysis of the progression of subjective symptoms over time revealed a more pronounced decrease in subjective symptoms in the FAV00A-treated group compared to the placebo group at days 3, 7 and 14 after initiation of treatment (Fig. 4). Specifically, we found that by day 14, the score had decreased from 15.7 ± 2.2 to 7.1 ± 0.5 in the FAV00A group compared to 15.4 ± 1.8 to 8.0 ± 1.3 in the placebo group. The mean group differences were -1.2 score points on day 7 (p<0.0001) and -0.9 score points each on day 3 (p=0.0065) and day 14 (p<0.0001). In addition, we found that the absence of all above mentioned subjective symptoms was more pronounced in the FAV00A-treated group on both day 7 and day 14 compared to the placebo control (Fig. 4A). We also found that the subjective symptoms that contributed most to the sum score (i.e., weakness, headache, increased perceived body temperature, sore throat, and cough) abated faster in the FAV00A-treated group compared to the placebo group (Fig. 4B-F). These results are also true when treated and non-treated patients suffering from different virus infections were compared on day 14 (data not shown). These findings suggest that FAV00A treatment has a positive outcome on clinically relevant subjective symptoms, such as feelings of weakness, headache, sore throat and coughing.

**Figure 4.**
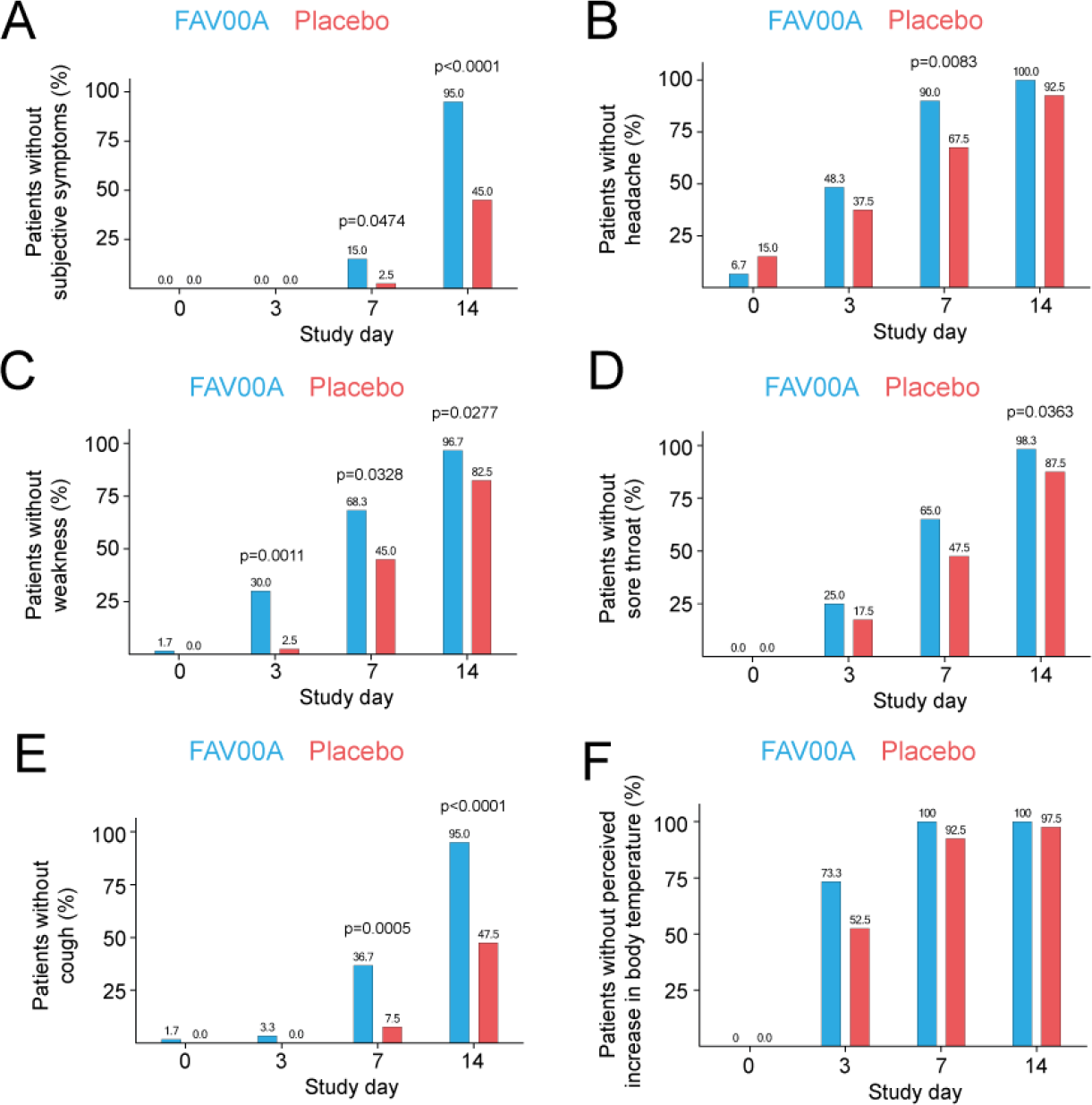
FAV00A treatment reduces subjective symptoms in patients with viral respiratory disease. A) Patients without subjective symptoms at different visit days (%). B) Patients without at different visit days (%). C) Patients without weakness at different visit days (%). D) Patients without sore throat at different visit days (%). F) Patients without cough at different visit days (%). E) Patients without elevated body temperature at different visit days (%). P values were determined by Fisher’s exact test.

### FAV00A treatment leads to faster patient recovery

Next, we analyzed the recovery time of FAV004-treated and placebo-treated patients in terms of resolution of symptoms and return to premorbid health status. In support of the above described observations, a complete recovery from symptoms associated with the viral respiratory infections was reported by 18.3% (n = 11) of the patients treated with FAV00A on day 7, while none in the placebo group had fully recovered on that day. At day 14, 93.3% (n = 56) of the FAV00A-treated patients had fully recovered, whereas only 32.5% (n = 13) of patients in the placebo group returned to normal health status. A significant improvement in the patient’s health status was reported by 60.0% (n = 36) of the FAV00A-treated patients compared to 15.0% (n = 6) of the placebo-treated patients on day 3 (p < 0.0001). FAV00A treatment led to a significant improvement in patient’s health at all three visit days after initiation of treatment (days 3, 7, 14, p < 0.0001 to p = 0.0018) when compared to the placebo group.

While FAV00A treatment generally led to faster patient recovery, 11 adverse events (AEs) occurred during the clinical trial. Of these, 4 AEs were reported by 4 patients (6.7%) treated with FAV00A (bitter taste in the mouth in 2 patients; heart burn and burning sensation in the throat in 1 patient each). The other 7 AEs in the FAV00A-treated patients were of mild intensity and resolved without additional therapy. Gastrointestinal side effects as reported by 4 patients are mentioned in the Amizon® Drug Product Label (Summary of Product Characteristics), which suggests that the AEs were related to Amizon®.

### FAV00A influenza A virus infection in cell culture

Previous *in vitro* experiments showed that FAV00A can inhibit IAV infection in normal human bronchial epithelial (NHBE) cultures (12). To confirm and extend these results, we incubated several human cells lines (A549, RD, Caco-2, or HepG2) with FAV00A and subsequently infected these cell lines with influenza A/WSN/1933 (H1N1) virus using MOIs that were optimized for each cell line. We found that FAV00A significantly affected WSN titers in A549 cells with an IC_50_ of 322 µM, in line with previous observations (Fig. 5A and 5E). Higher IC_50_ values were observed for the RD, Caco-2 and HepG2 cells (Fig. 5B, C, D and E). Selectivity indexes for FAV00A were >11 for A549, RD, and Caco-2 cells (Fig. 5E), but lower for HepG2 cells due FAV00A’s higher cytotoxicity in these cells (Fig. 5D and E). These data confirm that FAV00A can inhibit influenza A virus infections in a variety of human cell lines derived from lung, liver, colon and skeletal muscle tissues; however, its antiviral effects are strongly cell-type dependent, with respiratory tract derived cells such as NHBE (12) and A549 cells (Fig. 5A) being the best target cells for the anti-influenza A inhibitory activity of FAV00A.

**Figure 5:**
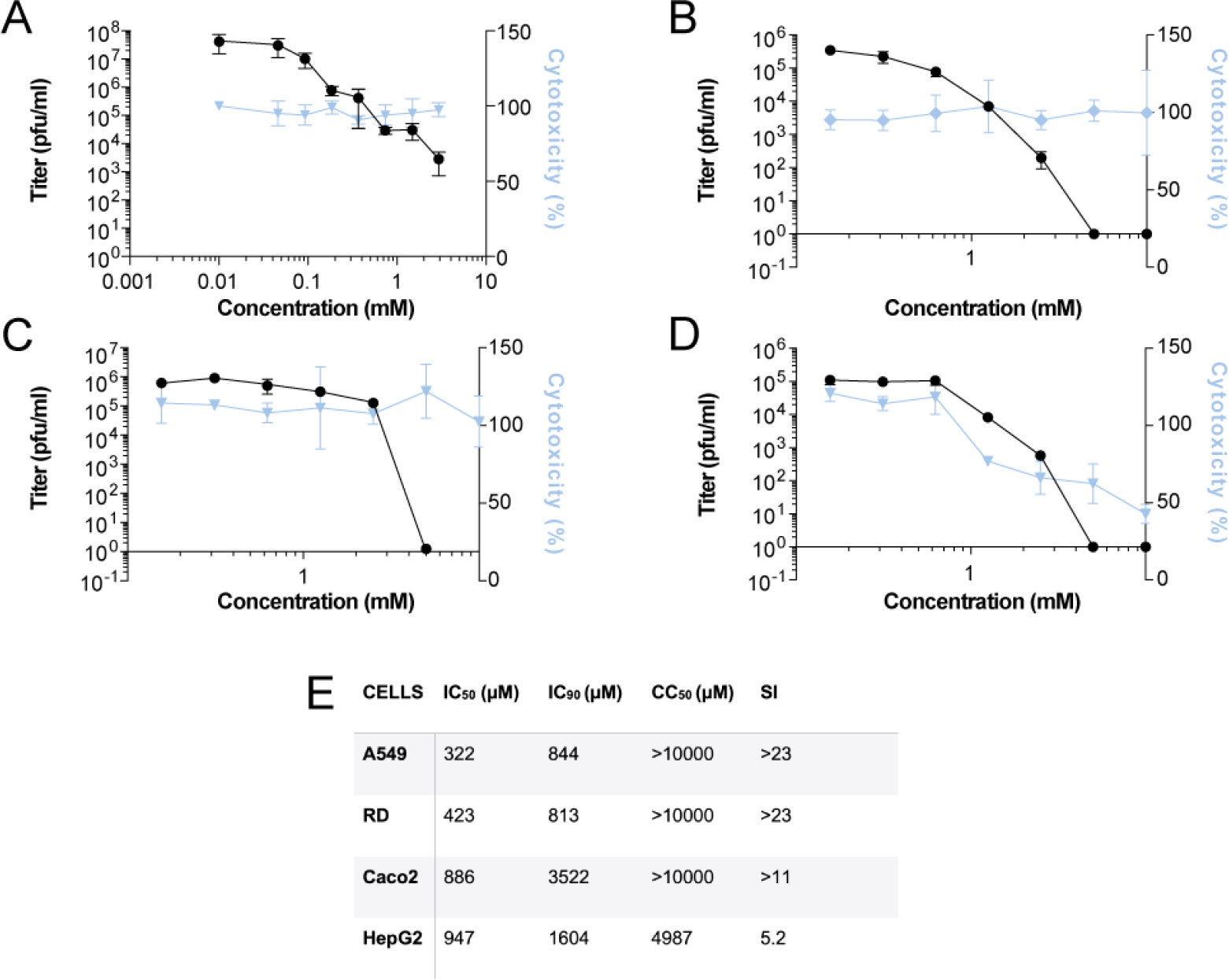
Inhibition of IAV infection and RNA synthesis by enisamium in cell culture. A) Effect of FAV00A on influenza A/WSN/1933 (H1N1) virus titers in A549 cells as determined by plaque assay (black line). Cytotoxicity (blue line) was determined in uninfected cells after 48 h of incubation with FAV00A. B) Effect of FAV00A on influenza A/WSN/1933 (H1N1) virus titers in RD cells, C) Caco-2 cells, and D) HepG2 cells as determined by micro-plaque assay (black line). Cytotoxicity (percentage of live cells, blue line) was determined in uninfected cells after 48 h of incubation with FAV00A. Data points represent mean and standard deviation of three independent FAV00A titrations and matching virus plaque experiments. E) Overview of IC_50_, IC_90_, CC_50_ and selectivity index (SI) values in different cell lines.

### FAV00A inhibits IAV RNA polymerase activity in cell culture

To test the mechanism of action for FAV00A, experiments were designed to study its activity on IAV RNA synthesis. For this purpose, an influenza minigenome assay was used as follows: HEK 293T cells were transfected with plasmids expressing the IAV RNA polymerase subunits PB1, PB2 and PA (pcDNA3-PB1, pcDNA3-PB2, pcDNA3-PA), the viral NP (pcDNA3-NP) and a vRNA template based on segment 5 (NP-encoding genome segment), all derived from the H1N1 WSN influenza virus (14, 15). After transfection, FAV00A was added to the cell culture medium at the concentrations indicated in Fig. 6A. Total cellular RNA was extracted 24 h post-transfection, and IAV replication (vRNA synthesis) activity was determined by radioactive primer extension analysis (16). FAV00A treatment significantly reduced the replication capacity of the viral RNA polymerase with an IC_50_ of 354 µM. To estimate the effect of FAV00A on host cell RNA synthesis, we transfected a plasmid expressing eGFP from a constitutively active CMV promoter and measured eGFP expression in the cell suspension of FAV00A treated cell cultures using GFP fluorescence. As a further control, we analyzed the effect of FAV00A treatment on the ribosomal RNA (rRNA) steady state level. No significant effect on 5S rRNA or GFP expression levels was observed in the presence of even higher concentrations (>2 mM) of FAV00A (Fig. 6A). Overall, these observations suggest that FAV00A directly affects influenza viral RNA polymerase activity in cell-based assays.

**Figure 6.**
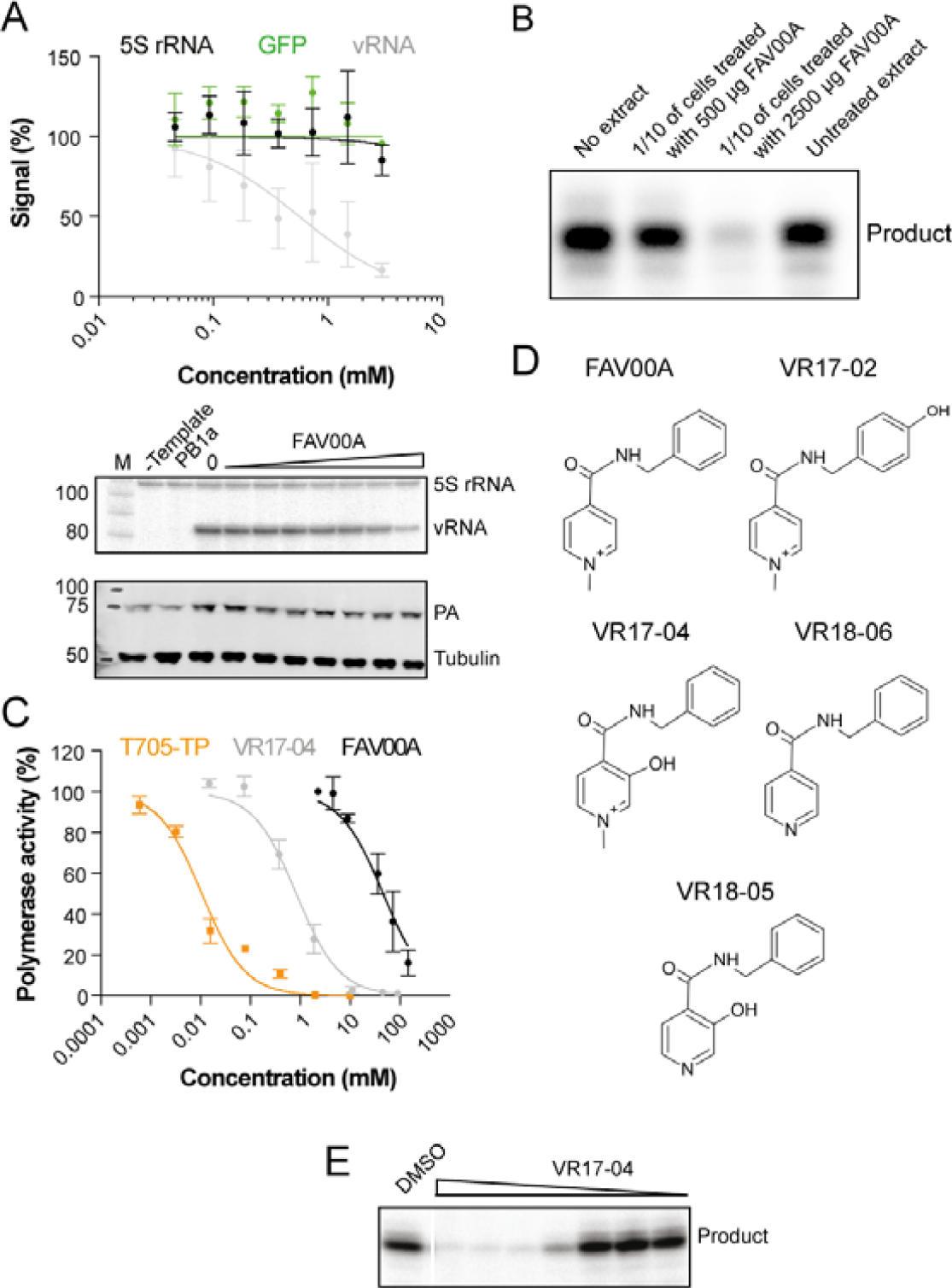
FAV00A is metabolized in humans and metabolite VR17-04 inhibits the viral RNA polymerase in vitro. A) Effect of FAV00A on the steady-state IAV vRNA, 5S rRNA and GFP levels with quantification shown in top graph. Levels of 5S rRNA and IAV vRNA were analyzed by primer extension (middle panel). PA and tubulin expression were analyzed by western blot (bottom panel). A mutant IAV RNA polymerase containing two amino acid substitutions in the PB1 active site (PB1a) was used as negative control. Data points represent mean and standard deviation of three independent FAV00A titrations and matching GFP measurements or primer extensions. B) Effect of extracts from A549 cells treated with FAV00A on IAV RNA polymerase activity in vitro. Five hundred or 2500 µg of FAV00A was added to A549 cells for 24 h. Next, cells were lysed and 1/10 of the lysate was added to in vitro polymerase assays. RNA polymerase products were analyzed by 20% denaturing PAGE. C) Quantification of the activity of the IAV RNA polymerase in vitro in the presence of FAV00A, VR17-04 or favipiravir triphosphate (T705-TP). Data points represent mean and standard deviation of three independent titrations in RNA polymerase assays. D) Phase I metabolites identified in human plasma samples. E) Activity of the IAV RNA polymerase in the presence of different VR17-04 concentrations.

### FAV00A weakly inhibits IAV RNA polymerase activity in vitro

To examine the effect of FAV00A on IAV RNA synthesis *in vitro*, we expressed protein A-tagged RNA polymerase from A/WSN/1933 (H1N1) virus in HEK 293T cells and purified the heterotrimeric TAP-tagged polymerase complex using IgG sepharose chromatography. We then performed *in vitro* RNA synthesis assays using a model 14 nucleotide (nt) vRNA template in the presence of different concentrations of FAV00A, or T-705 triphosphate (Fig. 6B) as a positive control. In these assays, FAV00A weakly inhibited IAV RNA polymerase activity with an IC_50_ of 46.3 mM, while T-705 triphosphate efficiently inhibited the RNA polymerase with mean IC_50_ of 0.011 mM (Fig. 6B). This suggests that FAV00A exerts an inhibitory effect on the IAV RNA polymerase activity *in vitro*, but its inhibitory activity is rather limited.

### FAV00A is metabolized in humans and metabolite VR17-04 inhibit IAV RNA polymerase activity

The weak inhibition of the IAV RNA polymerase activity by FAV00A is unlikely to be responsible for the obvious antiviral efficacy of FAV00A observed in cell culture and in patients. Based on the data obtained with FAV00A treatment in different cell types (Fig. 5), we hypothesized that a metabolite of FAV00A could be the actual inhibitor of the IAV RNA polymerase in cell culture. To test this hypothesis, we treated A549 cells with FAV00A for 24 h, lysed the cells and then performed IAV RNA polymerase activity assays in the presence of the cellular extract. We observed a strong, concentration-dependent inhibition of IAV RNA polymerase activity, which is consistent with the hypothesis that a metabolite of FAV00A inhibits influenza virus RNA synthesis during IAV infection (Fig. 6C).

To confirm that FAV00A is metabolized in humans, we analyzed human plasma samples from a phase I pharmacokinetic study using HPLC and MS/MS. This analysis revealed the presence of 4 phase I metabolites (Fig. 6D), of which two were hydroxylated (VR17-02 and VR17-04), one was demethylated (VR18-06), and one was both hydroxylated and demethylated (VR18-05). We also detected phase II metabolites that had been formed by glucuronidation or sulfate conjugation of phase I metabolites. Since glucuronidation or sulfate conjugation typically result in drug inactivation, we did not study the phase II metabolites further. In order to test if FAV00A phase I metabolites have a direct effect on IAV RNA polymerase activity *in vitro*, the phase I metabolites were synthesized. Only VR17-04 was sufficiently soluble in DMSO for *in vitro* testing and subsequently tested in *in vitro* IAV RNA polymerase assays containing purified IAV RNA polymerase, a 14-nt long vRNA, ApG, radiolabeled [a-^32^P]GTP, and all four unlabeled NTPs (14). Analysis of the reactions by 20% denaturing PAGE revealed a strong inhibition of viral RNA synthesis in the presence of the hydroxylated metabolite VR17-04 when compared to the DMSO control (Fig. 6E). Quantitation of the reactions showed that the human metabolite VR17-04 inhibited IAV RNA polymerase activity 55-fold stronger (IC_50_ of 0.84 mM) than FAV00A (IC_50_ of 46 mM) (Fig. 6C).

## Discussion

Infections with respiratory viruses create a huge burden on our health and economy, but currently only a limited number of antivirals are available against the viruses that cause them. Here, we investigated the efficacy and safety of FAV00A in patients aged between 18-60 years with clinically and virologically confirmed influenza and other viral respiratory infections. The clinical study demonstrated that treatment with FAV00A resulted in reduced virus shedding and a reduced mean number of days without routine activities in patients with viral respiratory infections, when compared to placebo. In addition, objective symptoms (fever, pharyngeal hyperemia) and subjective symptoms (weakness, headache, increased perceived body temperature, sore throat, and cough) were reduced and disappeared more quickly in patients receiving FAV00A treatment. These results suggest (i) a direct clinical effect of FAV00A on the outcome of viral respiratory infections in humans and (ii) that FAV00A treatment leads to faster patient recovery and a reduction of symptoms.

The use of concomitant symptomatic therapy (e.g., topical decongestants and expectorants) as well as the impact of the concomitant medication on the evaluation of the symptoms could affect clinical trial results. However, in the FAV00A-treated group, symptomatic drugs were taken less frequently and for shorter durations than in the placebo group, suggesting that an overestimation of the FAV00A effects due to concomitant symptomatic therapy can be excluded. In fact, it is tempting to suggest that FAV00A treatment led to reduced symptomatic therapy in the FAV00A-treated group. Moreover, in recent clinical trials with influenza virus neuraminidase inhibitors (e.g., oseltamivir), the evaluated symptoms such as fever, chills, headache, muscle ache, and cough as well as other clinical parameters return to routine activities, were similar to the symptoms and parameters evaluated in this clinical trial (17, 18). Therefore, the methods used to evaluate the efficacy of FAV00A for human respiratory virus infections and the results presented here should be reliable.

We also evaluated symptoms that are not characteristic for respiratory tract infections such as abnormal arterial blood pressure, abnormal auscultation findings of the heart, enlarged lymph nodes, and conjunctival infection. Most of these symptoms were indeed absent in the trial population or only present in a low percentage of the patients and of mild intensity. These non-characteristic symptoms were revealed in all or almost all patients on day 3 after initiation of treatment in both treatment groups, suggesting that these atypical symptoms did not contribute considerably to the outcome of the clinical trial. The patients included in this study had primarily mild to moderate respiratory disease. Thus, the efficacy of FAV00A against severe forms of respiratory disease or for high-risk groups of patients need to be confirmed. The results will also need to be confirmed in a double-blinded clinical trial, since the current study relied on blinded patients but unblinded investigators. While our study included predominantly patients infected with influenza viruses or co-infections between influenza viruses and adenovirus, the results suggest that FAV00A can be effective against other respiratory virus infections. Indeed, results from another recent study showed that FAV00A inhibits SARS-CoV-2 replication *in vitro* (19).

Analysis of serum samples from FAV00A-treated patients revealed a number of FAV00A metabolites, suggesting that not FAV00A, but one or more metabolites may be inhibiting respiratory virus infections. A previous study had suggested that FAV00A inhibits influenza virus RNA synthesis (12). Here, we confirmed the inhibitory effect of FAV00A on influenza virus RNA synthesis employing influenza virus minigenome experiments (Fig. 6). Moreover, using *in vitro* influenza virus RNA polymerase assays, we demonstrated that FAV00A can directly inhibit influenza virus replication and transcription and that one of its metabolites, VR17-04, is a more potent inhibitor of influenza virus RNA synthesis than FAV00A (Fig. 6). These findings are in line with the inhibitory effect of extracts from cultured human lung cells treated with FAV00A (Fig. 5); these observations might imply that FAV00A acts on the influenza virus RNA polymerase after hydroxylation to VR17-04.

In summary, we have performed a single-blinded clinical trial that shows that FAV00A treatment of patients with viral respiratory infections leads to faster patient recovery and reduced virus shedding when compared to the placebo control. Our results are supported by virus infections and viral replication *in vitro* studies on cells infected with IAV and with influenza viral RNA replication assays that suggest that FAV00A can directly inhibit influenza viral replication through a hydroxylated metabolite, VR17-04. This study thus advances our understanding of FAV00A and suggests that it could represent an alternative or additional treatment for current and emerging respiratory virus infections.

## Supporting information

CONSORT check list

## Data Availability

Clinical data is available from on ClincalTrials.gov under NCT04682444

## Acknowledgements

The authors thank Dr. Juergen Richt (Kansas State University, Manhattan, KS, US), Dr. Elena A. Govorkova (St. Jude Children Research Hospital, Memphis, TN, US), Dr. Peter Palese (Icahn School of Medicine at Mount Sinai (NYC, NY, US) and Adolfo Garcia-Sastre (Icahn School of Medicine at Mount Sinai, NYC, NY, US) for comments and advice.

## Transparency declarations

Aartjan te Velthuis is employed by the University of Cambridge, United Kingdom. The University of Cambridge received consulting fees for the experiments and analyses performed by Aartjan te Velthuis.

Megan Shaw was employed by the Icahn School of Medicine at Mount Sinai, New York, NY which received consulting fees for the experiments and analyses performed.

Andrew Mehle is employed by the University of Wisconsin – Madison, US. The University of Wisconsin – Madison received consulting fees for work performed for this study by Andrew Mehle.

Norbert Gmeinwieser received personal fees from Pharmalog Institut für klinische Forschung GmbH, subcontracted service CRO for writing the manuscript.

Holger Stammer received personal fees from Pharmalog Institut für klinische Forschung GmbH, subcontracted service CRO for review of the manuscript.

Jens Milde received personal fees from Pharmalog Institut für klinische Forschung GmbH, subcontracted service CRO for review of the manuscript.

Lutz Müller received personal fees from Dr. Regenold GmbH, subcontracted consultant for product development and for review of the manuscript.

Victor Margitich received personal fees from JSC Farmak, sponsor of the clinical trial and funding of the medical writing, for planning and coordinating the experiments and review of the manuscript.

