## Supplementary material for "Enisamium reduces influenza virus shedding and improves patient recovery by inhibiting viral RNA polymerase activity": CONSORT check list

**Items to include when reporting a randomized trial in a journal or conference abstract**

| **Item** | **Description** | **Reported on line number** |
| --- | --- | --- |
| Title | Identification of the study as randomized | 1-2 |
| Authors * | Contact details for the corresponding author | 3-4 |
| Trial design | Description of the trial design (e.g. parallel, cluster, non-inferiority) | 104-105 |
| Methods |  |  |
| Participants | Eligibility criteria for participants and the settings where the data were collected | 126-133 |
| Interventions | Interventions intended for each group | 133-134 |
| Objective | Specific objective or hypothesis | 104 |
| Outcome | Clearly defined primary outcome for this report | 144 ff. |
| Randomization | How participants were allocated to interventions | 133-134 |
| Blinding (masking) | Whether or not participants, care givers, and those assessing the outcomes were blinded to group assignment | 105 |
| Results |  |  |
| Numbers randomized | Number of participants randomized to each group | 279 |
| Recruitment | Trial status | 105 |
| Numbers analysed | Number of participants analysed in each group | 279-280 |
| Outcome | For the primary outcome, a result for each group and the estimated effect size and its precision | 299 ff. |
| Harms | Important adverse events or side effects | 368 ff. |
| Conclusions | General interpretation of the results | 449 ff. |
| Trial registration | Registration number and name of trial register |  |
| Funding | Source of funding | 513 |

**this item is specific to conference abstracts*
